# Mediation analysis in sibling designs: an application to the effect of prenatal antidepressant exposure on toddler depression mediated by gestational age at birth

**DOI:** 10.1101/2020.06.11.20128496

**Authors:** Mollie E. Wood, Espen Eilertsen, Eivind Ystrom, Hedvig Nordeng, Sonia Hernandez-Diaz

## Abstract

**Background:** Mediation analysis requires strong assumptions of no unmeasured confounding. Sibling designs offer a method for controlling confounding shared within families, but no previous research has done mediation analysis using sibling models.

**Methods:** We demonstrate the validity of the sibling mediation approach using simulation, and show its application using the example of prenatal antidepressant exposure and toddler anxiety and depression, with gestational age at birth as a mediator. We used data from the Norwegian Mother and Child Cohort Study, a cohort comprising 41% of births in Norway between 1999 and 2008 to identify 91,333 pregnancies, of which 25,776 were part of sibling groups.

**Results:** In simulations, sibling models were less biased than cohort models in cases where non-shared confounding was weaker than shared confounding, and when stronger non-shared confounding was controlled, but more biased otherwise. In the full cohort, the estimated mean difference in depression/anxiety scale z-scores for natural direct effects (NDE) were 0.31 (95% confidence interval 0.23 to 0.39) and 0.14 (95% CI 0.03 to 0.24), without and with adjustment for non-shared confounders, respectively. The natural indirect effect was 0.01 (95% CI 0.00 to 0.02) after adjustment. Adjustment for shared and non-shared confounding showed similar point estimates with wider confidence intervals (NDE 0.18, 95% CI −0.21 to 0.47; NIE −0.01, 95% CI −0.06 to 0.06).

**Conclusions:** Findings suggest that the modest association between prenatal antidepressant exposure and anxiety/depression is not mediated by gestational age and is likely explained by both shared confounders and non-shared confounders, and chance.

## Introduction

Causal inference research has codified theory and methods for estimating direct and indirect effects of an exposure on an outcome, through a particular pathway and through other means. These methods can provide insights on the pathways through which an exposure acts on an outcome, yielding mechanistic insights and suggesting possible interventions. However, the counterfactual framework for mediation analysis makes several strong assumptions if natural direct and indirect effects are to be identified including (i) no unmeasured confounding of the exposure-outcome association, (ii) no unmeasured confounding of the exposure-mediator association, (iii) no unmeasured confounding of the mediator-outcome association, and (iv) no unmeasured confounding of the mediator-outcome association that is affected by the exposure. Depending on the research question and data source, important confounders may be difficult or impossible to measure, or even unknown to the researcher.

Prenatal antidepressant exposure has been associated with an increased risk of neurodevelopmental outcomes in childhood including behavioral problems, milestone delays, and psychiatric diagnoses.(1–7) However, residual confounding is a serious challenge when interpreting these associations. Some studies have shown that adjustment for underlying maternal depression and/or anxiety reduces or eliminates these associations.(8–12) Prior studies have also linked prenatal antidepressant use to increased risks for shortened gestation.(13–18) Given that prematurity is strongly associated with poor neurodevelopmental outcomes,(19) understanding the mechanism by which antidepressant exposure may affect neurodevelopment requires appropriate consideration of this potential intermediate; however, adjustment for gestational age is complex.(20)

Mediation analysis provides a method for disentangling the direct effects of prenatal antidepressant exposure on child neurodevelopment from the potential indirect effects mediated through prematurity.(21,22) Studying whether there is an effect of antidepressants on neurodevelopment mediated through prematurity presents two major methodologic challenges. First, the maternal indications for the prescription of these treatments, mainly depression and anxiety, are themselves risk factors for both prematurity and neurodevelopmental problems in the infant.(8,23) Second, a naïve estimation of direct effects of antidepressant exposure on child behavior by controlling for gestational age at birth can result in biased estimates.(24)

We propose extending the sibling study design (25,26) for mediation analysis to control for shared familial confounding not only of the exposure-outcome association but of the exposure-mediator and mediator-outcome associations as well. While a previous study has examined mediators shared by siblings, with the goal of determining whether the sibling design may induce bias by inadvertently controlling for shared post-exposure variables,(27) no studies have examined using sibling methods to control unmeasured shared confounders when estimating direct and indirect effects of exposure. The aim of this paper was to evaluate the performance of this approach in the presence of unmeasured shared and non-shared (by siblings) confounders using simulations; and apply this combination of methods to help clarify the mechanisms by which prenatal antidepressant exposure is associated with toddler anxiety.

## Material and Methods

### Counterfactual Effect Definitions

We use A_ij_, Y_ij_, and M_ij_ to denote the exposure, outcome, and mediator, respectively, for sibling *j* (*j=*1,2) in family *i*. U_i_ is a shared confounder in family *i*. For example, if A_ij_ is prenatal exposure to antidepressants, Y_ij_ is a measure of toddler anxiety/depression symptoms, and M_ij_ is gestational age at birth, U_i_ may represent some combination of parental genetic risk and familial environment. L_ij_ is a non-shared confounder, such as maternal depressive symptoms during different pregnancies. A possible causal model is outlined in Figure 1.

**Figure 1.**
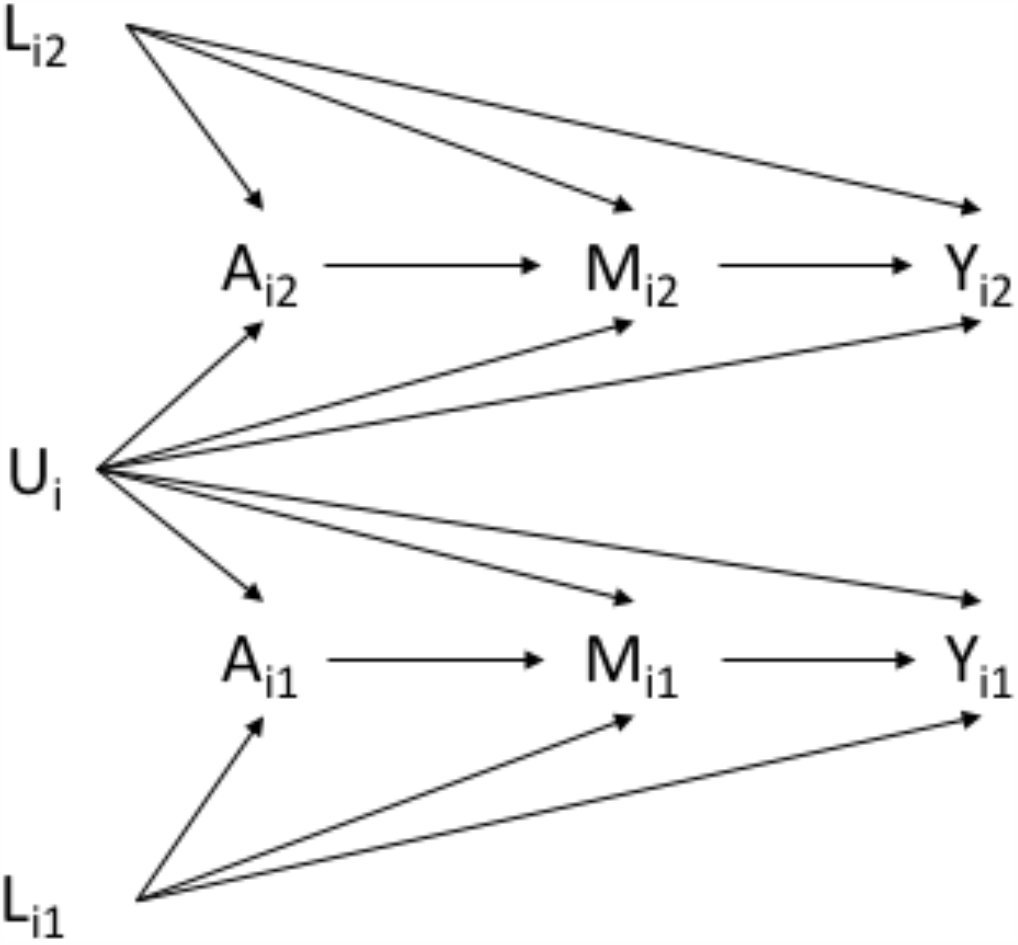
Possible causal model for an exposure *A*, mediator *M*, outcome *Y*, non-shared confounder *L*, and shared confounder *U*, for sibling *j* (*j=*1,2) in family *i*. In the example using data from the Norwegian Mother and Child Cohort Study, exposure is antidepressant use in pregnancy, mediator is gestational age at birth, outcome is toddler neurodevelopment, non-shared confounders are maternal depressive or anxiety symptoms, age, parity marital status, smoking, alcohol use, and other medication use measured in pregnancy *j*, and shared confounders are any heritable risk factors such as genetics or family environment that are stable in family *i*.

### Regression models for mediation analysis

We adapted existing parametric methods for estimation of the total effect (TE), controlled direct effect (CDE), natural direct effect (NDE), and natural indirect effect (NIE) by extending methods described by Vanderweele and Valeri.(22) The standard approach to mediation analysis is to fit two regression models: one for the mediator, conditional on exposure and confounders, and a second for the outcome, conditional on exposure, mediator, and confounders, and allowing for an exposure-mediator interaction as in equation (1) and (2), below.

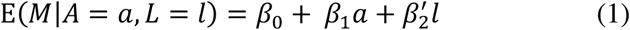

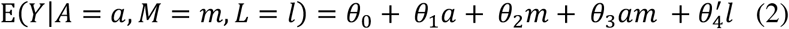

Coefficients from the regression model are used to construct the relevant effects:

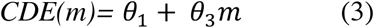

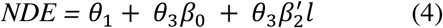

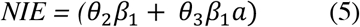

In our adapted approach, we fit sibling models for the mediator (equation 6) where E(*M*_*ij*_ |*A*_*ij*_,*Ā* _j_,*L*_*j*_) is the conditional mean of *M*_*ij*_, given individual exposure *A* _*ij*_, mean sibling exposure, *Ā* _*i*_ and non-shared confounders *L*_*j*_ and for the outcome (equation 7) where 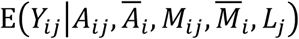 is the conditional mean of *Y*_*ij*_ given individual exposure *A*_*ij*_ and mediator *M*_*ij*_, mean sibling exposure *Ā*_*i*_ and mediator 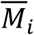, and non-shared confounders *L*_*j*_. This approach conditions on shared confounders U_ij_ by design.

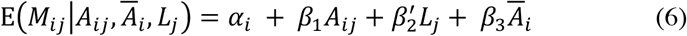

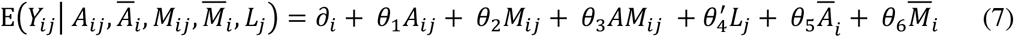

We then extract the relevant coefficients from these parametric models, as in the standard approach, to construct the effects of interest.

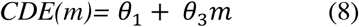

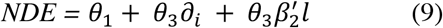

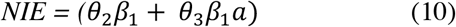

The controlled direct effect is the difference in outcome when comparing A=1 to A=0, conditional on L=l and U=u for a fixed level of M (equation 8); in our example, we could define this as the effect of antidepressant exposure on toddler anxiety and depressive symptoms, if we could intervene to set gestational age at birth to 280 days for all pregnancies, and conditioned on important shared and non-shared confounders. The natural direct effect, given in (9), is the effect of antidepressant exposure on internalizing behavior if gestational age at birth were distributed as in the unexposed. The natural indirect effect (10) is the effect on the outcome after setting the mediator to have the distribution it would have had in the exposed, versus in the unexposed.(22)

### Simulation Study

Data for the simulation were generated for pairs of observations, as shown in Figure 1. The non-shared confounder, L, was an independent random variable with prevalence of 50%, A is a dichotomous exposure conditional on L, M is a continuous mediator conditional on A and L, U is an unmeasured shared confounder of A-M, AY, and M-Y, and Y is a continuous outcome conditional on A, M, and L:

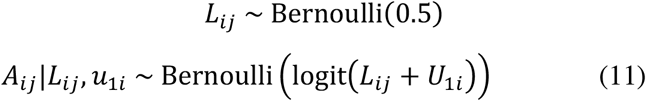

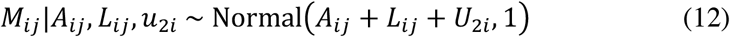

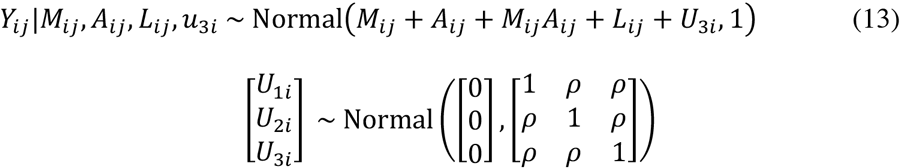

We fixed the A-M path to 1.0, the M-Y path to 2.0, and the A-Y path to 5.0, to reflect a scenario with both a direct and indirect effect of A on Y. We also generated an interaction term between A and M, and a nonshared confounder, L. Within this framework, we varied the strength of the effect of L on A, M, and Y (0.0, 1.0, 5.0) and the correlation P between siblings (0.0, 0.2, 0.6). We generated data for 100,000 pairs and then collected and pooled the results for all combinations of P and effects of L. Note that this approach generates a cohort of pairs; i.e., in our simulated cohort, all observations are siblings. Further details on the simulation are included in the supplementary material.

For each regression model, we fit the model first without consideration for the nested structure of the data (referred to hereafter as a “cohort” approach), and then fit between-within models in the same model to control shared effects within families by including the sibling-pair mean exposure and mediator (hereafter referred to as “sibling model”). Thus, we estimated quantities for the total effect, controlled direct effect, natural direct effect, and natural indirect effect. We then repeated the data generation and analysis procedure 1000 times and calculated the % bias for each scenario as [(observed-truth)/observed]*100.

### Practical Example: prenatal antidepressant exposure and anxiety/depression symptoms in toddlers

We used data from the Norwegian Mother and Child Cohort Study (MoBa), a questionnaire-based prospective birth cohort comprising 41% of pregnancies in Norway between 1999 and 2008, that advanced to at least 18 weeks gestation and were seen at a municipal or county health station for prenatal care. Data for this study are from the quality-assured version 9 of the dataset, linked to the Medical Birth Register of Norway (MBRN).(28) The MoBa study has received a license from the Norwegian Data Inspectorate and approval from the Regional Committee for Medical Research Ethics, and all participants in MoBa gave written consent to participation. Details on study inclusion and exclusion are outlined in Figure 2, and differences between the sibling and non-sibling sample are detailed in supplemental table 1.

**Figure 2.**
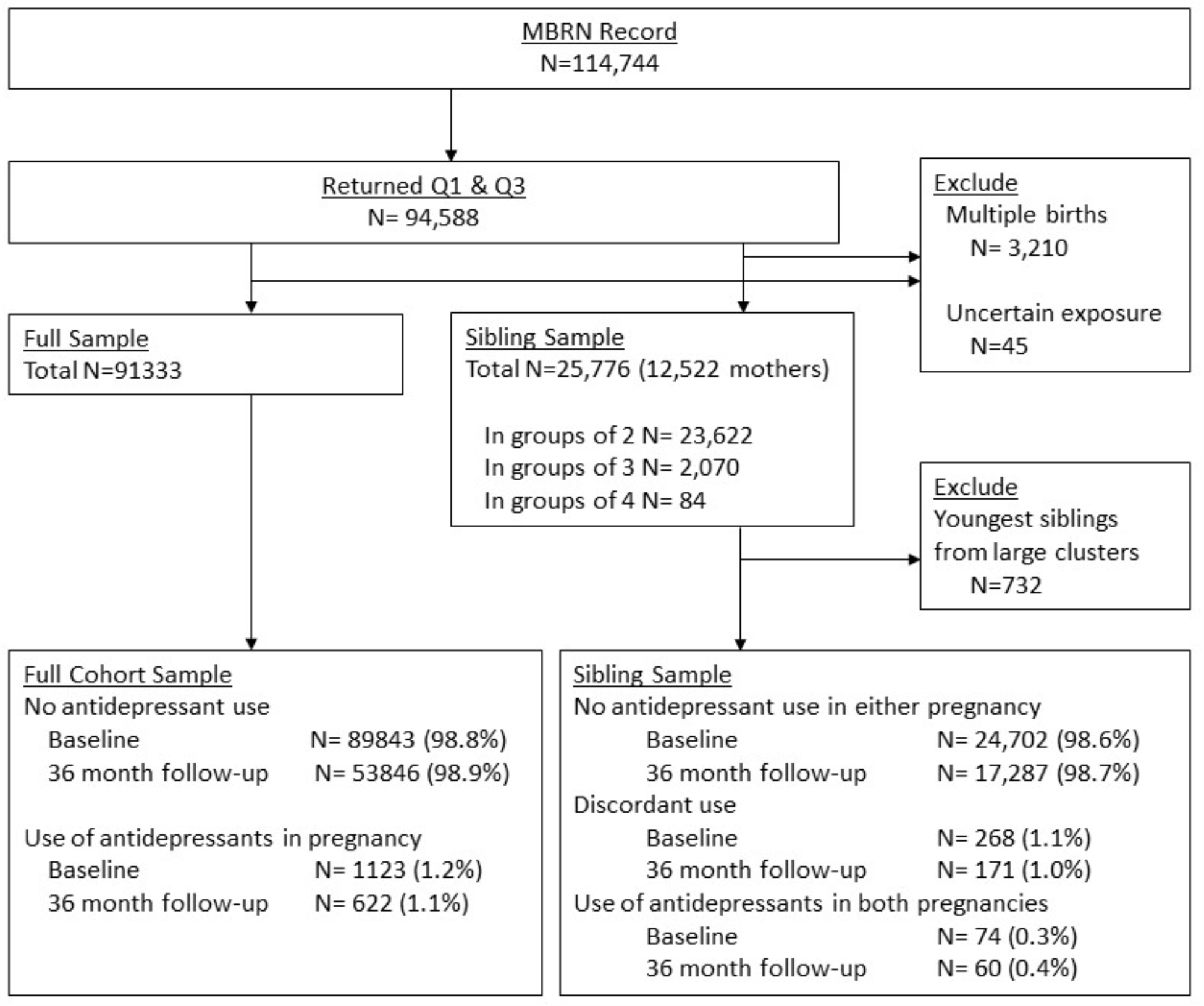
Study inclusion flow chart.

### Exposure

Exposure to antidepressants was ascertained via self-report on Questionnaire 1 (Q1, given between gestational weeks 17 and 30) and Questionnaire 3 (Q3, given after gestational week 30). Women reported the name of each drug taken and indicated the timing of exposure in 4-week intervals. Drugs were classified according to their ATC codes:(29) all antidepressants (ATC: N06A), SSRIs (ATC: N06AB), and non-SSRI antidepressants [including monoamine oxidase inhibitors (N06AF), non-selective monoamine reuptake inhibitors (N06AB), monoamine oxidase A inhibitors (N06AG) and other antidepressants (N06AX)]. Our exposure in these analyses was use of antidepressants at any time during pregnancy. As a sensitivity analysis, we examined exposure that occurred in the second or third trimester, as this definition better reflects persistent antidepressant use in pregnancy (results in supplemental figure 1).

### Mediator

Gestational age at birth was extracted from the Medical Birth Register of Norway.

### Outcome

Toddler Anxiety/Depression was assessed using an abbreviated version of the Child Behavior Checklist (CBCL)(30), a parent-reported outcome measure that has been validated in a Norwegian sample,(31) with higher positive scores indicating more problems. More details on the items included can be found in supplemental table 1.

### Potential confounders

We included maternal age, parity, marital status, smoking during pregnancy, alcohol use in pregnancy, concomitant use of medications (i.e. analgesics, other psychotropics), and current symptoms of depression and anxiety in early pregnancy as potential confounders. Maternal symptoms of depression and anxiety were assessed using a validated short version of the Hopkins Symptom Checklist-25 (SCL-25), a screening tool used to assess the severity of depression and anxiety during pregnancy; the SCL covers symptoms occurring in the two weeks prior to filling out the questionnaire.(32)

### Statistical Analysis

To estimate the natural direct effect (NDE) and natural indirect effect (NIE) of prenatal antidepressant exposure on internalizing behavior, we first fit a linear model for the mediator, with gestational age at birth as the dependent variable and antidepressant exposure as the main explanatory variable, while controlling for measured confounders. Next, we fit a linear model for the outcome, in which internalizing behavior was the dependent variable, and antidepressant exposure, gestational age, an interaction term for exposure and mediator, and measured non-shared covariates were included as predictors. For both the mediator and outcome models, we first fit models in the full cohort (including mothers who participated only once, as well as those who participated multiple times), then in the sibling sample, which comprised only women who participated two or more times. All models were fit with and without control for measured non-shared confounders (maternal age, smoking, alcohol use, marital status, education, parity, depressive and anxiety symptoms, lifetime history of depression, concomitant medications, and pain diagnoses). Models in the sibling cohort were additionally adjusted for the sibling-pair mean exposure and mediator. Following model fitting, we used model coefficients to calculate the CDE, NDE, and NIE. The within-pair coefficient, which is generally interpreted as the causal effect of exposure after adjusting for shared and non-shared confounding, was used as the “treatment effect” coefficient in models including the sibling-pair mean exposure and/or mediator. The total effect (TE) is the sum of the NDE and NIE. Confidence intervals were estimated using 1000 bootstrapped samples. All analyses were carried out using R/RStudio, using the *lm, lme4*, and *boot* packages.

## Results

### Simulation results

The results of the simulation study for a dichotomous exposure and continuous mediator and outcome models are presented in Figure 3. Results, expressed as percent bias, are divided into the CDE (top panel), NDE (middle panel), and NIE (lower panel), with no shared confounding on the left, modest (P=0.2) shared confounding in the center, and strong (P=0.6) shared confounding on the right. Non-shared confounding increases from left to right within each panel, and estimates controlling for non-shared confounding by L are shown in solid black, while estimates where non-shared confounding by L is uncontrolled are shown by unfilled points and dashed lines. Estimates from sibling models were less biased than cohort models for all scenarios except those in which the sibling correlation was weak or absent *and* non-shared confounding was strong and uncontrolled. For all scenarios, the natural direct effects (NDE) and natural indirect effects (NIE) were more sensitive to bias than the controlled direct effect (CDE).

**Figure 3.**
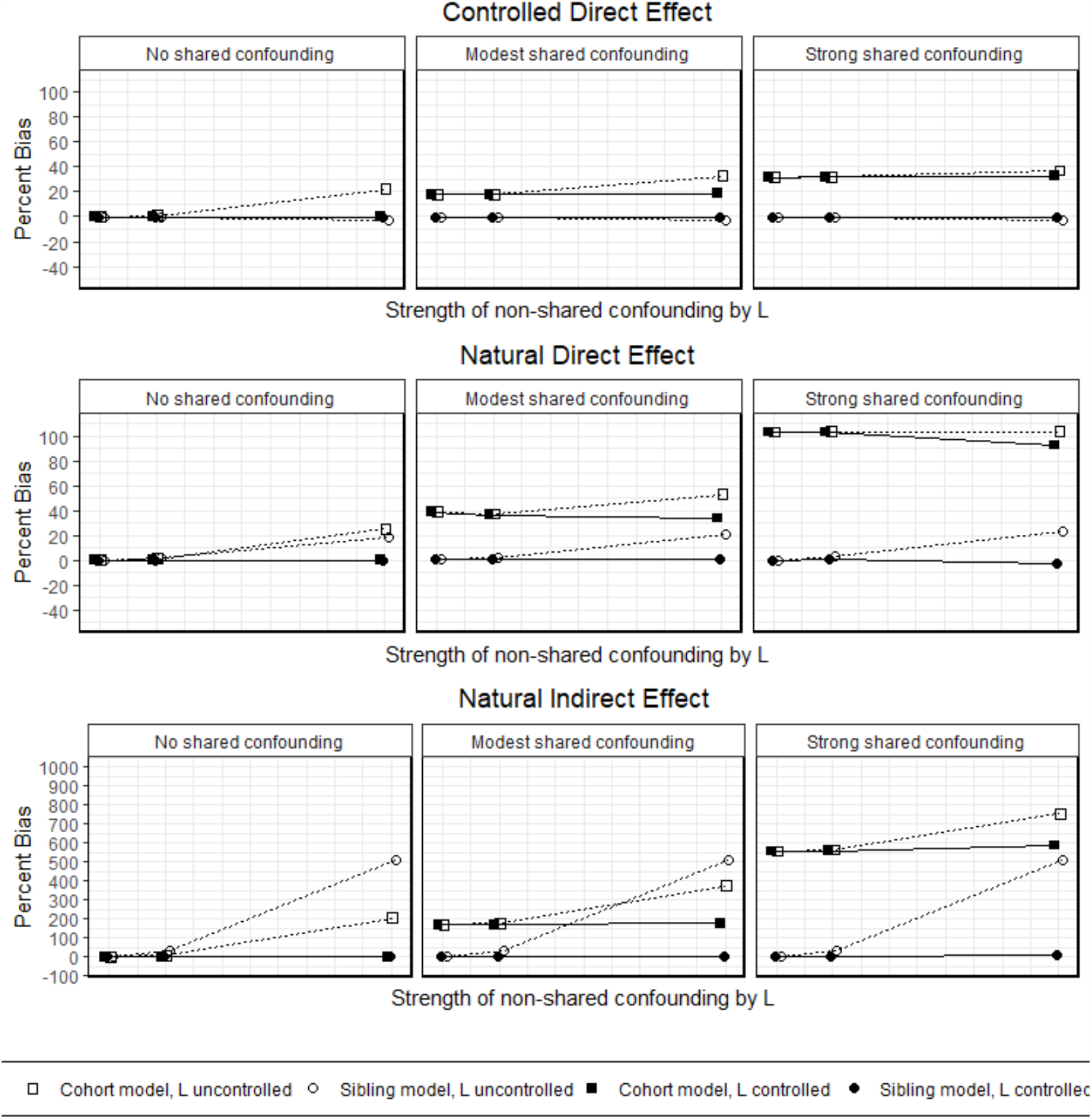
Results of simulation study comparing the performance of cohort and sibling models in different confounding scenarios, for a linear mediator and linear outcome. Percent bias {([truth-observed]/truth)*100} is presented for total effect (TE), controlled direct effect (CDE), natural direct effect (NDE), and natural indirect effect (NIE) for sibling and cohort models under the following scenarios: (A) no shared confounding, L uncontrolled, (B) no shared confounding, L controlled, (C) modest shared confounding, L uncontrolled, (D) modest shared confounding, L controlled, (E) strong shared confounding, L uncontrolled, and (F) strong shared confounding, L controlled.

### Results from the MoBa

Characteristics of full cohort are presented in Table 1. Of the 25,044 eligible women participating in the MoBa with two pregnancies, 208 (0.8%) reported antidepressant use during at least one of their pregnancies. Antidepressants used were mainly SSRIs (77%). Women who used antidepressants were more often primiparous, less likely to be married or living with a partner, more likely to have a lifetime history of major depression and/or higher symptoms of depression or anxiety during their pregnancy; they also reported higher rates of smoking and alcohol consumption as well as more frequent use of analgesics and benzodiazepines during pregnancy. Exposed children were more likely to be female, which may be related to the selection of the sibling sample, as the sibling sample includes a higher proportion of girls than the main MoBa sample (supplemental table 2). A comparison of the within-sibling pair characteristics is included in supplemental table 3.

**Table 1.** Baseline sample characteristics by antidepressant exposure status for the Norwegian Mother and Child Cohort Study sample

|  | Unexposed<sup>1</sup><br>(n=90206) | Exposed<br>(n=1127) |
| --- | --- | --- |
| Maternal age | 30.2(4.6) | 30.1(5.6) |
| Parity >0, % | 55 | 47 |
| Married or cohabitating, % | 97 | 89 |
| Used SSRI antidepressants in pregnancy, % | 0 | 79 |
| Used non-SSRI antidepressants in pregnancy, % | 0 | 24 |
| History of major depressive symptoms, % | 6 | 44 |
| Depressive/anxiety symptoms in early pregnancy (z score) | -0.01(0.99) | 0.93(1.31) |
| Depressive/anxiety symptoms in late pregnancy (z score) | -0.02(0.98) | 0.94(1.35) |
| Smoked cigarettes during pregnancy, % | 12 | 27 |
| Any alcohol use in pregnancy, % | 16 | 20 |
| Use of other medications in pregnancy |  |  |
| Benzodiazepines, % | <1 | 10 |
| Anti-epileptic drugs, % | <1 | 3 |
| Opioids, % | 2 | 5 |
| Paracetamol, % | 43 | 52 |
| NSAIDs, % | 7 | 12 |
| Child characteristics |  |  |
| Gestational age at birth (days) | 279.7(11.9) | 278.0(11.9) |
| Female, % | 49 | 51 |
| Birth weight (grams) | 3609(545) | 3556(544) |

Values are means (SD) or percentages.
<sup>1</sup>Use at any time during pregnancy.

Results for the mediation analysis to decompose the effect of antidepressant exposure on toddler anxiety and depression symptoms into direct and indirect effects are presented in Table 2 and Figure 4. In the main cohort models, our initial unadjusted effect estimate suggested an effect of antidepressant exposure partially mediated by gestational age at birth, which attenuated but did not disappear after covariate adjustment for observed confounders. Carrying out analyses in the smaller sibling cohort produced further attenuation of estimates; however, most of the decrease occurred in the natural direct effect, while a small effect through gestational age remained. After adjusting for measured confounders, the only remaining observed association between prenatal antidepressant exposure and toddler anxiety/depression was through the effect of antidepressants on gestational age at birth. Results were similar when exposure was defined as use in the second or third trimester, albeit with wider confidence intervals due to a smaller number of exposed (figure S1).

**Table 2.** Estimates of direct and indirect effects of the association between prenatal antidepressant exposure and child anxiety/depression symptoms at 36 months, and the proportion of antidepressant exposure-related behavior that is mediated through gestational age, with and without adjustment for shared and non-shared confounders

|  |  | Total Effect |  | Controlled Direct Effect<sup>1</sup> |  | Natural Direct Effect |  | Natural Indirect Effect |  |
| --- | --- | --- | --- | --- | --- | --- | --- | --- | --- |
|  |  | D<sup>2</sup> | 95% CI | D | 95% CI | D | 95% CI | D | 95% CI |
| Full cohort sample (N=54,468) |  |  |  |  |  |  |  |  |  |
|  | No adjustment | 0.32 | 0.24, 0.40 | 0.31 | 0.23, 0.39 | 0.31 | 0.23, 0.39 | 0.01 | 0.00, 0.03 |
|  | Adjusted for non-shared confounders<sup>3</sup> | 0.15 | 0.04,0.25 | 0.14 | 0.03, 0.24 | 0.14 | 0.03, 0.24 | 0.01 | 0.00, 0.02 |
| Sibling sample (N=17,518) |  |  |  |  |  |  |  |  |  |
|  | No adjustment | 0.26 | 0.08, 0.43 | 0.21 | 0.03, 0.38 | 0.21 | 0.03, 0.38 | 0.05 | -0.01, 0.14 |
|  | Adjusted for only non-shared confounders<sup>3</sup> | 0.09 | -0.09, 0.26 | 0.04 | -0.13, 0.22 | 0.04 | -0.13, 0.22 | 0.04 | -0.01, 0.12 |
|  | Adjusted for only shared confounders<sup>4</sup> | 0.26 | -0.02, 0.57 | 0.26 | -0.02, 0.56 | 0.26 | -0.02, 0.56 | 0.00 | -0.04, 0.05 |
|  | Adjusted for shared and nonshared<br>confounders | 0.18 | -0.20, 0.50 | 0.18 | -0.21, 0.47 | 0.18 | -0.21, 0.47 | -0.01 | -0.06, 0.06 |

1. Effect of antidepressant exposure on internalizing behavior, fixing gestational age at birth to 280 days.
2. Mean difference in anxiety/depression symptoms, in standard deviation units, associated with prenatal exposure to antidepressants, compared to no exposure; positive coefficients indicate increases in internalizing behavior.
3. Models adjusted for maternal age, education, marital status, parity, concomitant medication use, smoking, alcohol use, symptoms of depression/anxiety in pregnancy, and lifetime history of major depression.
4. "Adjusted for shared confounders" indicates models conditioned on mean exposure and mediator in sibling pair, thereby adjusting for all measured and unmeasured confounders shared within sibling pair.

**Figure 4.**
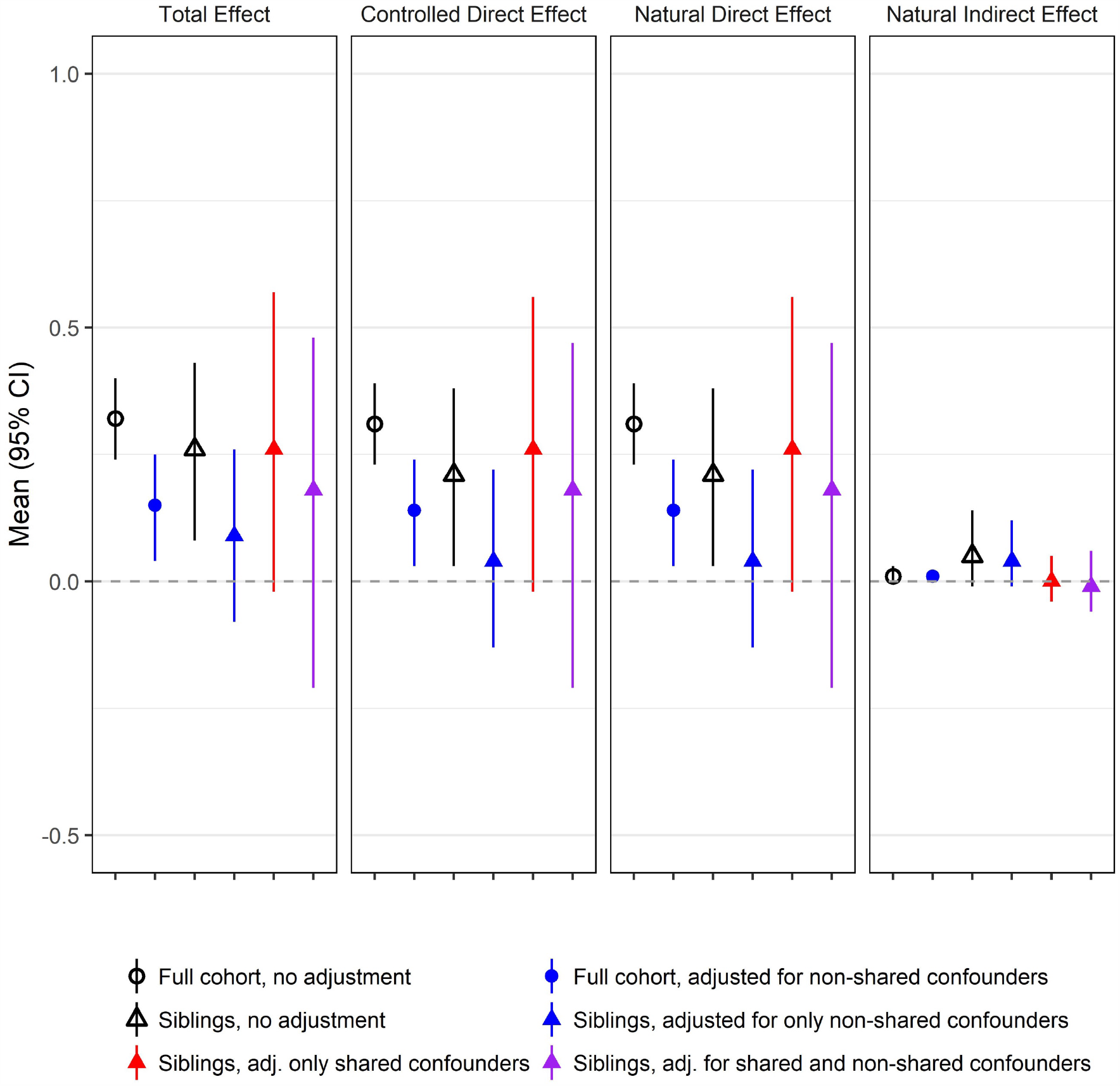
Estimates of direct and indirect effects (mediated through gestational age at birth) of the association between prenatal antidepressant exposure and child anxiety/depression symptoms at 36 months, and the proportion of antidepressant exposure-related neurodevelopment that is mediated through gestational age, with and without adjustment for non-shared confounders

In sibling models adjusting for sibling mean exposure and mediator, we observed a similar initial estimate of total effect in models unadjusted for non-shared confounders. In contrast with the cohort models, sibling models showed no evidence of an indirect effect of antidepressant exposure on internalizing behavior though gestational age. Additional adjustment for non-shared confounders resulted in attenuation of the TE and NDE, although not as much as in cohort models. Importantly, confidence intervals were substantially wider for sibling models.

## Discussion

Our study resulted in two main findings. First, based on simulations, when important confounders are either shared within families or non-shared and measured, mediation analysis in sibling studies returns unbiased results, compared with the same analysis performed without stratification on family. Therefore, for the scenarios we considered (e.g., no measurement error and correlated exposures), including a sibling analysis may be a useful sensitivity analysis to address residual unmeasured shared/familial confounding.

Second, in the application, we found that gestational age at birth does not appear to be an important mediator for the effect of prenatal antidepressant exposure on internalizing behavior at 36 months of age. Considering the similar unadjusted estimates obtained in the cohort and the sibling designs and the substantial attenuation of the estimate after adjustment for measured confounders, findings suggest that most of the association between prenatal antidepressant exposure and neurodevelopmental outcomes is due to confounding, particularly by factors specific to each pregnancy. In addition, non-shared confounding appears more important for the direct antidepressant-anxiety association, while shared/familial confounding was more important for the indirect effect through gestational age. Limiting consideration to the effect estimates, we might suggest that failing to carry out this analysis using a sibling design would have resulted in overestimating the natural indirect effect through gestational age, possibly due to inadequate control of unmeasured, shared confounders of the mediator-outcome path; however, the wide confidence intervals around these estimates mean that a more likely explanation is random error and lack of precision, and correspondingly unstable effect estimates.

Much discussion in the causal inference literature concerns the necessity of well-defined interventions,(33) and gestational age at birth may be conceptually problematic as a mediator in this context, as there are multiple ways to set the mediator to a particular outcome. Gestational age at birth is not especially amenable to intervention in any real-world setting. Vanderweele demonstrated that the natural indirect effect is interpretable as a particular kind of mediated effect, but that the direct effect (e.g. the effect of antidepressants on toddler anxiety not mediated through gestational age) estimates both a true direct effect and also an effect mediated through other forms of the mediator.(34) We propose that this kind of analysis is still of interest when investigating the mechanisms by which prenatal drug exposure acts on later child development, as evidenced by many studies that (incorrectly) simply adjust for gestational age, but we recognize that opinions diverge on this matter.(35)

Sibling study samples may have different characteristics than the general birthing population. In our study, we found that women who participated multiple times were on average older, more likely to be married, were less often taking antidepressants, and had less severe symptoms of depression and anxiety during pregnancy. In addition, sibling studies come with their own set of assumptions. The most salient of these is outlined in Frisell et al (2012)(36): namely, that a causal interpretation of the within-sibling effect estimate requires the sibling correlation in confounders is stronger than the sibling correlation in exposures. This assumption is not formally testable in data and so relies strongly on subject-area knowledge. For the current problem, in which we attempt to disentangle the associations between antidepressant exposure *in utero* and toddler anxiety into those occurring indirectly through gestational age at birth, and where the underlying confounding is likely strongly correlated between siblings, using a between-within sibling approach is likely less biased than a traditional cohort model. Our results may not be generalizable to exposures and outcomes with different confounding characteristics. In addition, sibling designs assume no carryover effects from the first to the second sibling.(37) In exposure-discordant sibling pairs, 61% of the exposed vs. 40% of the unexposed were the oldest sibling in the group (supplemental table 2), and it is possible that experiences from the earlier pregnancy do inform behaviors in subsequent pregnancies. However, estimates from sensitivity analyses comparing results with and without adjustment for birth order were not substantially different (results not shown), suggesting that carryover effects are not a major source of bias in this study. Finally, we did not consider other potential sources of selection bias occurring at multiple occasions during the study, including selection on multiparity (when moving from the full cohort to the sibling sample) and selection on loss to follow up (when only including observations present at the 36-month assessment).

It is important to note that when considering depression as a confounder of the associations between antidepressant exposure, gestational age, and neurodevelopment, we often refer to both a *chronic* confounder (e.g., genetic liability) and an *acute* confounder (e.g., active depressive symptoms). By definition, the chronic confounder does not change over time, while the acute one often does. Therefore, which estimate we trust depends on which confounding scenario we believe: a cautious researcher would be well advised to consider both the cohort and sibling estimate as a means of putting empirical bounds around the possible causal effect of interest, and should additionally make use of existing sensitivity analysis methods to ascertain the possible impact of unmeasured confounding.(38)

## Conclusion

Disentangling direct and indirect effects of prenatal exposures may yield critical insights into the mechanisms by which exposures affect outcomes. The results of our study suggest that in some cases, the addition of sibling analyses to existing sensitivity analyses for unmeasured confounding in mediation analysis can yield valuable insights into the role of confounding and the plausibility of the major assumptions of this approach.

This work is supported by the National Institutes of Health (2T32HL098048-11; Wood), a European Research Council Starting Grant (grant number 639377; Nordeng, Ystrom), the Norwegian Research Council (grant number 262177; Eilertsen, Ystrom), and the Faculty of Mathematics and Natural Sciences, University of Oslo, Norway (Wood).

## Data Availability

Data from the MoBa can be accessed by application to the Norwegian Institute of Public Health. Code for reproducing the simulations in this manuscript are included in the supplementary material.

## Acknowledgements

The Norwegian Mother and Child Cohort Study is supported by the Norwegian Ministry of Health and Care Services and the Ministry of Education and Research, National Institutes of Health/National Institute of Neurological Disorders and Stroke (grant no.1 UO1 NS 047537-01 and grant no.2 UO1 NS 047537-06A1). We are grateful to all the participating families in Norway who take part in this on-going cohort study.

The authors would like to thank Dr. Olga Basso for comments on an early version of the manuscript.

## Abbreviations

AD: antidepressants
CDE: controlled direct effect
CI: confidence interval
MoBa: Norwegian Mother and Child Cohort Study
NDE: natural direct effect
NIE: natural indirect effect
SSRI: selective serotonin reuptake inhibitors

## Supplemental Material

**Table S1.** Items included in Child Behavior Checklist (CBCL) Anxious/Depressed behavior subscale. Response options are “Not true”, “Somewhat or sometimes true”, “Very often true”

| Subdomain | Items |
| --- | --- |
| Emotionality reactive | • Disturbed by any change in routine |
| Anxious / depressed | • Clings to adult or too dependent<br>• Gets too upset when separated from parents<br>• Too fearful or anxious<br>• Nervous, high strung or tense<br>• Unhappy, sad or depressed<br>• Self-conscious or easily embarrassed<br>• Feelings are easily hurt |
| Somatic complaints | • Does not eat well<br>• Stomach aches or cramps (without medical cause)<br>• Vomiting, throwing up (without medical cause) |

**Table S2.** Comparison of sibling and non-sibling sample

|  | Non-sibling sample<br>(n=65557) | Sibling sample<br>(n=25776)<sup>1</sup> |
| --- | --- | --- |
| Maternal age* | 30.24(4.72) | 30.04(4.14) |
| Parity >0, % | 52 | 63 |
| Married or cohabitating, % | 96 | 98 |
| Used SSRI antidepressants in pregnancy, % | 1 | 1 |
| Used non-SSRI antidepressants in pregnancy, % | 0 | 0 |
| History of major depressive symptoms, % | 6 | 5 |
| Depressive/anxiety symptoms in early pregnancy (z score) | 0.08(0.97) | -0.08(0.89) |
| Depressive/anxiety symptoms in late pregnancy (z score) | 0.07(0.99) | -0.10(0.90) |
| Smoked cigarettes during pregnancy, % | 14 | 8 |
| Any alcohol use in pregnancy, % | 17 | 15 |
| Other medications used in pregnancy |  |  |
| Benzodiazepines, % | 1 | 0 |
| Anti-epileptic drugs, % | 0 | 0 |
| Opioids, % | 2 | 2 |
| Paracetamol, % | 43 | 43 |
| NSAIDs, % | 7 | 6 |
| Child Characteristics |  |  |
| Gestational age at birth (days) | 279.6(12.0) | 279.9(11.4) |
| Female, % | 49 | 49 |
| Birth weight (grams) | 3597(551) | 3638(526) |

Values are means(SD) or percentages.
1. Includes 732 pregnancies later excluded from sample (sibling number 3 or 4 from women who participated more than 2 times)

**Table S3.**
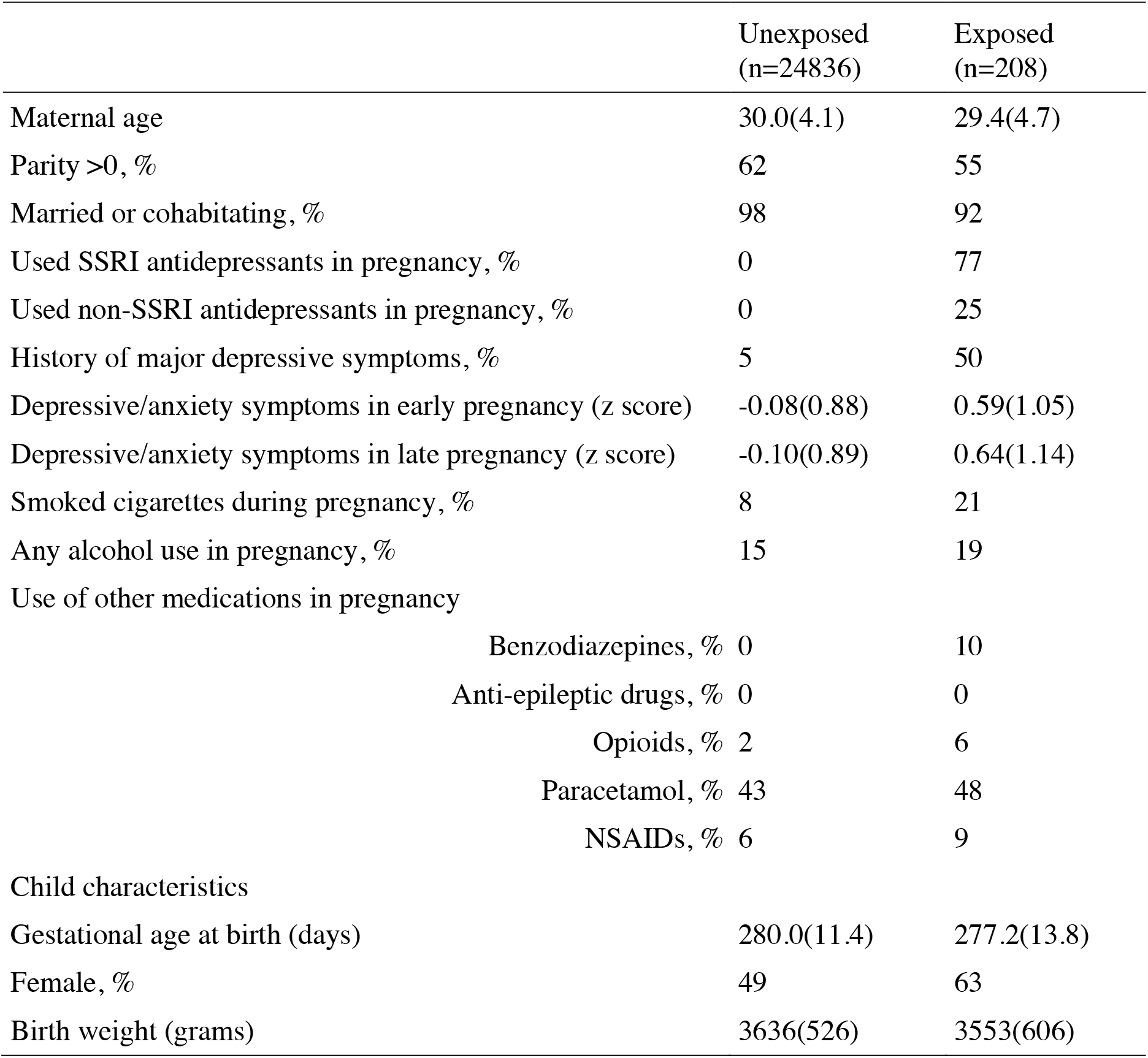
Comparison of exposed versus unexposed pregnancies in the sibling sample

**Table S4.** Comparison of differences between sibling pairs for exposure concordant and discordant siblings

|  | Discordant exposure |  | Concordant exposure<br>(both exposed) |  | Concordant exposure<br>(both unexposed) |  |
| --- | --- | --- | --- | --- | --- | --- |
|  | Exposed<br>(n=134) | Unexposed<br>(n=134) | Sibling 1<br>(n=37) | Sibling 2<br>(n=37) | Sibling 1<br>(n=12351) | Sibling 2<br>(n=12351) |
| Maternal age | 28.87(4.49) | 29.43(4.62) | 29.08(4.66) | 31.76(4.76) | 28.64(3.89) | 31.31(3.92) |
| Parity >0, % | 53 | 70 | 19 | 100 | 23 | 100 |
| Married or cohabitating, % | 90 | 95 | 94 | 97 | 97 | 99 |
| Used SSRI antidepressants in pregnancy, % | 77 | 0 | 76 | 81 | 0 | 0 |
| Used non-SSRI antidepressants in pregnancy, % | 24 | 0 | 30 | 24 | 0 | 0 |
| History of major depressive symptoms, % | 46 | 34 | 57 | 59 | 5 | 5 |
| Depressive/anxiety symptoms in early pregnancy (z score) | 0.69(1.02) | 0.66(1.14) | 0.32(0.98) | 0.54(1.19) | -0.06(0.87) | -0.11(0.89) |
| Depressive/anxiety symptoms in late pregnancy (z score) | 0.68(1.19) | 0.73(1.30) | 0.70(1.02) | 0.40(1.07) | -0.04(0.92) | -0.17(0.86) |
| Smoked cigarettes during pregnancy, % | 26 | 16 | 14 | 11 | 10 | 6 |
| Any alcohol use in pregnancy, % | 16 | 14 | 31 | 17 | 17 | 13 |
| Benzodiazepines, % | 9 | 4 | 16 | 5 | 0 | 0 |
| Anti-epileptic drugs, % | 1 | 1 | 0 | 0 | 0 | 0 |
| Opioids, % | 7 | 4 | 3 | 8 | 2 | 2 |
| Paracetamol, % | 50 | 54 | 46 | 43 | 40 | 46 |
| NSAIDs, % | 10 | 10 | 8 | 8 | 6 | 5 |
| Child Characteristics |  |  |  |  |  |  |
|  | Exposed<br>(n=134) | Unexposed<br>(n=134) | Sibling 1<br>(n=37) | Sibling 2<br>(n=37) | Sibling 1<br>(n=12351) | Sibling 2<br>(n=12351) |
| Oldest sibling, % | 61 | 39 | 100 | 0 | 100 | 0 |
| Gestational age at birth (days) | 277.0(15.0) | 277.1(13.8) | 278.3(13.5) | 276.9(8.9) | 280.0(12.2) | 280.0(10.5) |
| Female, % | 62 | 44 | 68 | 59 | 49 | 48 |
| Birth weight (grams) | 3591(661) | 3603(630) | 3463(531) | 3506(449) | 3571(536) | 3703(506) |

Values are means(SD) or percentages.

**Table S5.** Estimates of direct and indirect effects of the association between late pregnancy antidepressant exposure and child anxiety/depression symptoms at 36 months with and without adjustment for non-shared confounders

|  |  | Total Effect |  | Controlled Direct Effect<sup>1</sup> |  | Natural Direct Effect |  | Natural Indirect Effect |  |
| --- | --- | --- | --- | --- | --- | --- | --- | --- | --- |
|  |  | D<sup>2</sup> | 95% CI | D | 95% CI | D | 95% CI | D | 95% CI |
| Full cohort |  |  |  |  |  |  |  |  |  |
|  | No adjustment | 0.40 | 0.18, 0.42 | 0.27 | 0.15, 0.39 | 0.27 | 0.15, 0.40 | 0.03 | -0.01, 0.06 |
|  | Adjusted for non-shared confounders<sup>3</sup> | 0.13 | -0.03, 0.28 | 0.09 | -0.07, 0.25 | 0.09 | -0.07, 0.25 | 0.04 | -0.01, 0.08 |
| Sibling sample |  |  |  |  |  |  |  |  |  |
|  | No adjustment | 0.23 | 0.03, 0.45 | 0.20 | -0.04, 0.43 | 0.20 | -0.04, 0.43 | 0.03 | -0.07, 0.17 |
|  | Adjusted for only non-shared confounders | 0.09 | -0.13, 0.31 | 0.07 | -0.17, 0.28 | 0.07 | -0.17, 0.28 | 0.02 | -0.07, 0.14 |
|  | Adjusted for only shared confounders | 0.24 | -0.26, 0.76 | 0.22 | -0.27, 0.77 | 0.22 | -0.27, 0.76 | 0.02 | -0.13, 0.11 |
|  | Adjusted for shared and nonshared confounders | 0.32 | -0.52, 0.89 | 0.31 | -0.40, 0.90 | 0.31 | -0.17, 0.28 | 0.01 | -0.22, 0.09 |

1. Effect of antidepressant exposure on internalizing behavior, fixing gestational age at birth to 280 days.
2. Mean difference in anxiety/depression symptoms, in standard deviation units, associated with prenatal exposure to antidepressants, compared to no exposure; positive coefficients indicate increases in internalizing behavior.
3. Models adjusted for age, parity, concomitant medication use, smoking, alcohol use, symptoms of depression/anxiety in pregnancy, and lifetime history of major depression.

**Figure S1.**
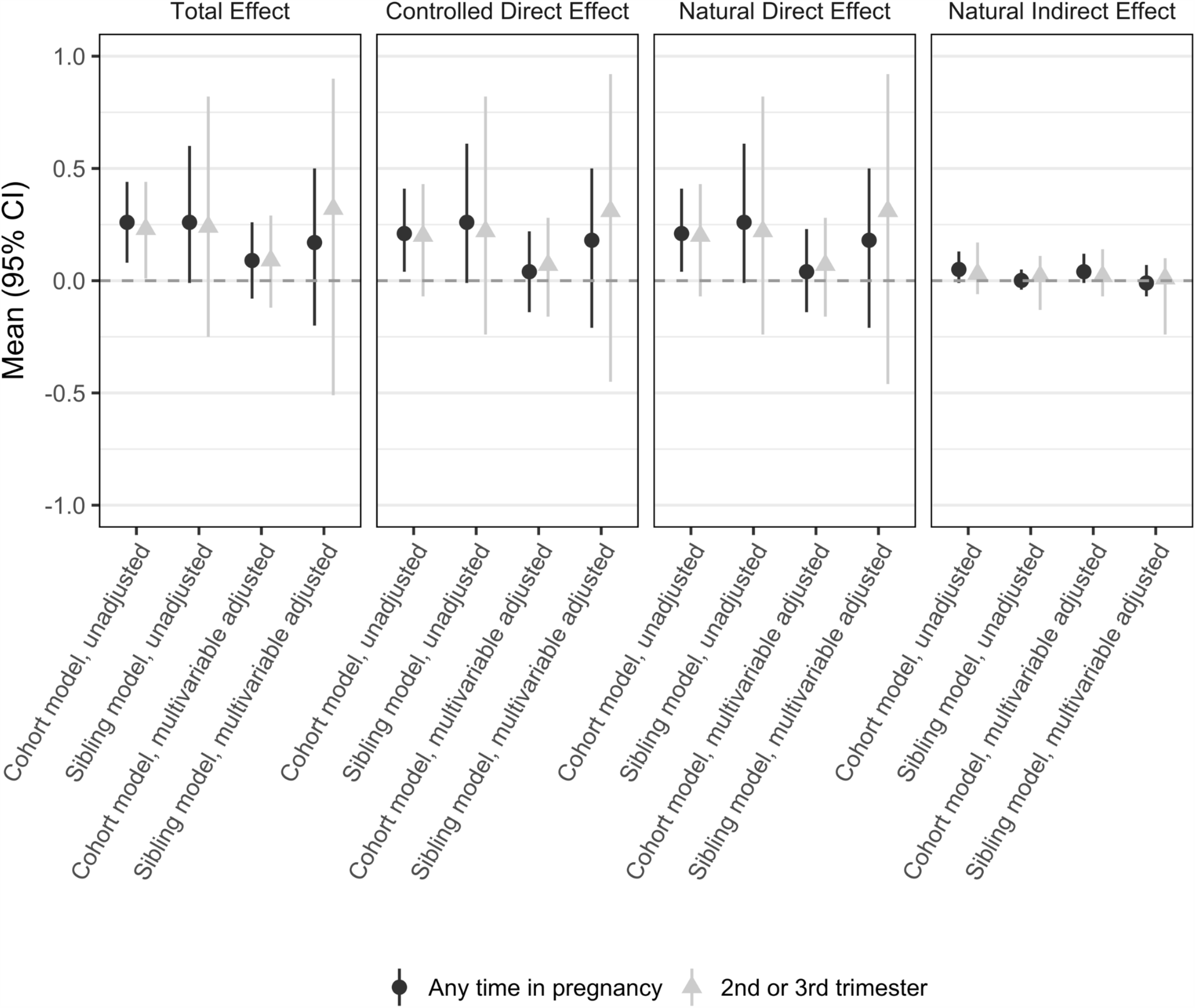
Alternative exposure definition

Appendix. R code for simulations

~~~
rm(list = ls())
library(MASS)
library(dplyr)
library(tidyr)
library(lme4)
library(survival)
library(psych)
J = 100000 # Number of sibling pairs
#######################################################
## FOR CONTINUOUS OUTCOME Y ##
#######################################################
#to manipulate “sharedness” of confounding, adjust sd_eta and r_eta
#for no confounding, set all except diagonal of r_eta to 0; set la/lm/ly to 0
# Y,M,A,L #0 for L because L is unshared
sd_eta = diag(c(5,5,5,0)) #sd of residuals for shared component (55% correlation) correlation of
residuals
 #A M Y L
r_eta = matrix(c(1.0, 0.6, 0.6, 0.0, #A
 0.6, 1.0, 0.6, 0.0, #M
 0.6, 0.6, 1.0, 0.0, #Y
 0.0, 0.0, 0.0, 0.0), 4, 4, byrow = T) #L: 0 because L is unshared
vc_eta = sd_eta%*%r_eta%*%t(sd_eta)
am = c(1.0, 1.0) #effect of exposure on mediator
my = c(2.0, 2.0) #effect of mediator on outcome
ay = c(5.0, 5.0) #effect of exposure on outcome
la = c(0.5, 0.5) #effect of confounder on exposure
lm = c(3.0, 3.0) #effect of confounder on mediator
ly = c(2.0, 2.0) #effect of confounder on outcome
z = matrix(NA, J, 10)
colnames(z) = c(“y1”, “y2”, “m1”, “m2”, “a1”, “a2”, “i1”, “i2”,”l1”, “l2”)
for(j in 1:J) {
 eta = mvrnorm(1, c(0, 0, 0, 0), vc_eta)
 ls = eta[4] + rlogis(2)
 l= ifelse(ls > 0,1,0)
 as = l*la + eta[3] + rlogis(2)
 a = ifelse(as > 0, 1, 0)
 m = am*a + l*lm + eta[2] + rnorm(2)
 i= a*m
 y = my*m + ay*a + l*ly + 0.5*i + eta[1] + rnorm(2)
 z[j, ] = c(y, m, a, i,l)
}
d = data.frame(z, pair = 1:J)
dll = d %>% gather(var, z, -pair) %>%
 mutate(sib = substr(var, 2, 2),
 var = substr(var, 1, 1)) %>%
 arrange(pair, sib)
dl = dll %>%
 spread(var, z) %>%
 group_by(pair) %>%
 mutate(mean.m = mean(m),
 mean.a = mean(a))
#testing for bias in model coefficients
#can add or remove l to assess effect of controlling unshared confounder
######################################################
# cohort model
# Model for M
fit11 = lm(m∼ a + l, dl)
# MOdel for Y
fit21 = lm(y∼ m + a + i + l, dl)
summary(fit11)
summary(fit21)
cohort = c(coef(fit11)[2], coef(fit21)[2:3])
names(cohort) = c(“am”, “my”, “ay”)
# sibling model
# Model for M
fit12 = lmer(m∼ a + mean.a + l + (1|pair), dl)
# Model for Y
fit22 = lmer(y ∼ m + a + mean.a + mean.m + l + i + (1|pair), dl)
summary(fit12)
summary(fit22)
sibling = c(fixef(fit12)[2], fixef(fit22)[2:3])
names(sibling) = c(“am”, “my”, “ay”)
cohort
sibling
######################################################
mediation_cohort <- lm(m ∼ a + l, dl)
mediation_sibs <-lmer(m ∼ a + mean.a + l + (1|pair), dl)
beta_c_0 <-mediation_cohort$coef[1]
beta_c_a <-mediation_cohort$coef[2]
beta_c_l <-mediation_cohort$coef[3]
beta_s_0 <-fixef(mediation_sibs)[1]
beta_s_a <-fixef(mediation_sibs)[2]
beta_s_l <-fixef(mediation_sibs)[4]
outcome_cohort <-lm(y ∼ a + m + i + l, dl)
outcome_sibs <-lmer(y ∼ a + m + i + l + mean.a + mean.m +(1|pair), dl)
theta_c_a <-outcome_cohort$coef[2]
theta_s_a <-fixef(outcome_sibs)[2]
theta_c_m <-outcome_cohort$coef[3]
theta_s_m <-fixef(outcome_sibs)[3]
theta_c_i <-outcome_cohort$coef[4]
theta_s_i <-fixef(outcome_sibs)[4]
#fix CDE at sample mean
CDE_cohort <- (theta_c_a + theta_c_i*1.99)
CDE_sibs <- (theta_s_a + theta_s_i*1.99)
#marginal effects by fixing confounder at sample mean
NDE_cohort <-(theta_c_a + theta_c_i*beta_c_0 + theta_c_i*beta_c_a + theta_c_i*beta_c_l*0.5) NDE_sibs <- (theta_s_a + theta_s_i*beta_s_0 + theta_s_i*beta_s_a + theta_s_i*beta_s_l*0.5) NIE_cohort <- (theta_c_m*beta_c_a + theta_c_i*beta_c_a*1)
NIE_sibs <- (theta_s_m*beta_s_a + theta_s_i*beta_s_a*1)
TE_cohort <-NDE_cohort + NIE_cohort
TE_sibs <-NDE_sibs + NIE_sibs
PM_cohort <- (NIE_cohort/TE_cohort)*100
PM_sibs <- (NIE_sibs/TE_sibs)*100
~~~

